# Vitamin D Supplementation is Associated with Slower Epigenetic Aging

**DOI:** 10.1101/2021.12.10.21267597

**Authors:** Valentin Max Vetter, Yasmine Sommerer, Christian Humberto Kalies, Dominik Spira, Lars Bertram, Ilja Demuth

## Abstract

Adverse effects of low vitamin D level on mortality and morbidity are controversially discussed. Especially older people are at risk for vitamin D deficiency and therefore exposed to its potentially harmful influence on the aging process. A way of measuring differences in the biological age is through DNA methylation age (DNAm age) and its deviation from chronological age, DNAm age acceleration (DNAmAA). We previously reported on an association between vitamin D deficiency and higher 7-CpG DNAmAA in participants of the Berlin Aging Study II (BASE-II).

In this study, we employ a quasi-interventional study design to assess the relationship between DNAmAA of five epigenetic clocks and vitamin D supplementation. Longitudinal data were available for 1,036 participants of BASE-II that were reexamined on average 7.4 years later in the GendAge study (mean age at follow-up: 75.6 years, SD = 3.8 years, age range: 64.9 – 94.1 years, 51.9 % female). DNAmAA was estimated with the 7-CpG clock, Horvath’s clock, Hannum’s clock, PhenoAge and GrimAge. Methylation data were obtained through methylation-sensitive single nucleotide primer extension (MS-SNuPE) or Illumina’s Infinium “MethylationEPIC” array.

Vitamin D deficient participants who chose to start vitamin D supplementation after baseline examination showed a 2.6 year lower 7-CpG DNAmAA (p=0.011) and 1.3-year lower Horvath DNAmAA (p=0.042) compared to untreated and vitamin D deficient participants. DNAmAA did not statistically differ between participants with successfully treated vitamin D deficiency and healthy controls (p>0.16).

Therefore, we conclude that intake of vitamin D supplement is associated with lower DNAmAA in participants with vitamin D deficiency. Additionally, our findings suggest that sufficient vitamin D supplementation can compensate and potentially reverse the increase in 7-CpG DNAmAA that we found in in vitamin D deficient participants.

## Introduction

The health impact of vitamin D deficiency and benefits of its supplementation are subject to an ongoing controversial discussion [1]. Estimations suggest that 1 billion people worldwide have a serum vitamin D level below 50 nmol/L, the widely accepted cut-off for vitamin D deficiency [2]. Risk of vitamin D deficiency is particularly high for older adults because of their lower capability for cutaneous synthetization and less exposure to sunlight (homebound, institutionalized) [3]. Schöttker and colleagues found a constant decrease of on average 3 nmol/L serum vitamin D for every 10 years of age in a large population-based cohort (n=9,940, age range: 50-74 years) and therefore proposed vitamin D as a marker of aging [4].

The consequences of vitamin D deficiency are not fully understood. Prolonged and severe vitamin D deficiency as cause of rickets in children [5] and osteomalacia in adults [6] is uncontested. However, its role in prevention of falls [7-9] and fractures [2, 10-18] remains unclear. Besides its potential effect on musculoskeletal health, associations with numerous diseases such as respiratory tract infections [19, 20], depressive symptoms [21], rheumatoid arthritis [22, 23], multiple sclerosis [24], colorectal cancer [25, 26], breast cancer [27] and mortality [28] were reported, as well. These findings, however, are controversially discussed [29] and whether vitamin D deficiency is cause or result of these diseases is still unclear [30].

Considering the wide range of diseases associated with or promoted by vitamin D deficiency, a better understanding of its role in the process of healthy (biological) aging has far-reaching individual and socio-economic implications. Several biomarkers of aging are available that allow objectivation of biological age [31, 32]. The latest and often considered most promising [32] marker is the DNA methylation age (DNAm age) and its deviation from chronological age, DNAm age acceleration (DNAmAA). Different versions of epigenetic clocks that are used for its estimation are available. They differ mostly in number and location of analyzed cytosine-phosphate-guanin (CpG) sites as well as the domain of aging they represent best [33]. For most of these clocks the association between DNAm age and mortality (reviewed in [34]) and morbidity (reviewed in [32, 33]) is well documented. However, studies that examine the relationship between epigenetic age and vitamin D status are limited and inconclusive [4, 35, 36].

In this study we make use of two-wave longitudinal data on 1,036 older adults of the Berlin Aging Study II (BASE-II) that were reexamined on average 7.4 years later as part of the GendAge study. We previously reported on a cross-sectional association between vitamin D deficiency and high 7-CpG DNAmAA in this cohort at baseline [36]. We now repeated this analysis in the same participants at follow-up examination and compared it with DNAmAA obtained from four other epigenetic clocks, including GrimAge and PhenoAge [36]. Additionally, we employed a quasi-interventional (non-randomized) analysis scheme [37] to compare vitamin D deficient participants that chose to start vitamin D supplementation during the follow-up period with participants with “untreated” vitamin D deficiency at both time-points. Subsequently, we selected participants with sufficient vitamin D level to analyze DNAmAA in “treated” participants in comparison with “healthy” controls. We hypothesized, that the treatment of vitamin D deficiency would be associated with slower epigenetic aging and treated participants would not differ from participants without any signs for vitamin D deficiency.

## Methods

### Berlin Aging Study II (BASE-II) and GendAge Study

The Berlin Aging Study II (BASE-II) cohort consists of a convenience sample of 1,671 residents (age range: 60-85 years) of the greater metropolitan area of Berlin, Germany [38, 39] that was first examined between 2009 and 2014 (baseline, T0). The younger age group of BASE-II (n=500, age range: 20-37 years) has not been considered in this study.

On average 7.4 years later, 1,083 participants of the older group were re-examined as part of the GendAge study (follow-up, T1) [40]. Seventeen additional participants that were not assessed in the baseline examination were included at follow-up, resulting in a total dataset of 1,100 cases at follow-up.

All participants gave written informed consent. The medical assessments at baseline and follow-up were conducted in accordance with the Declaration of Helsinki and approved by the Ethics Committee of the Charité – Universitätsmedizin Berlin (approval numbers EA2/029/09 and EA2/144/16) and were registered in the German Clinical Trials Registry as DRKS00009277 and DRKS00016157.

### DNA Methylation Age (DNAm age)

We employed five different epigenetic clocks in this study. The 7-CpG clock [41] incorporates seven cytosine-phosphate-guanine (CpG) sites that were chosen to be most informative on chronological age and feasible to be measured by single nucleotide primer extension (SNuPE) [41-44]. Briefly, genomic DNA was isolated from whole blood samples and subsequently bisulfite converted. The segments of interest were amplified by a multiplex polymerase chain reaction (mPCR) and a single nucleotide primer extension (SNuPE) was performed.

Horvath’s clock [45], Hannum’s clock [46], PhenoAge [47] and GrimAge [48] were estimated from DNA methylation data that was obtained from the same DNA samples via Illumina’s “Infinium MethylationEPIC” array. Loading and quality control (QC) of the DNAm data was performed with the R package bigmelon [49] using default settings as described previously [50]. Probes that showed ≥1% of samples with a detection p-value of 0.05 or a bead count below three in >5% of samples were excluded from all analyses. Samples with a bisulfite conversion efficiency below 80% (estimated by the *bscon* function) and outliers (identified by the *outlyx* and *pcout* function [51]) were removed. After elimination of the outliers, the samples were reloaded and normalized with the *dasen* function. A root-mean-square deviation (RMSD) of ≥ 0.1 in DNAm-values after normalization, as determined by the *qual* function, led to exclusion of the sample from the dataset and loading and normalization were repeated with the new set of samples. Methylation age was calculated according to the manual of Steve Horvath’s website (https://horvath.genetics.ucla.edu/html/dnamage/). Of the 513 CpGs of Levine’s clock and 71 CpGs of Hannum’s clock, 512 CpGs and 64 CpGs were available in this dataset. For a more detailed description of methods, please refer to ref. [52].

### DNA Methylation Age Acceleration (DNAmAA)

The deviation of DNAm age from chronological age, DNAm age acceleration (DNAmAA), was calculated as residuals of a linear regression analysis of DNAm age on chronological age and cell counts (neutrophils, monocytes, lymphocytes, eosinophils) [53]. Cell counts were measured by flow-cytometry in an accredited standard laboratory (LaborBerlin, Berlin, Germany).

### Vitamin D

Vitamin D levels were measured as 25-hydroxycholecalciferol (25-OHD) in blood serum samples that were taken during the same blood draw as the whole blood samples used for DNA isolation. A standard laboratory (LaborBerlin, Berlin, Germany) determined the serum vitamin D level through chemiluminescence immunoassays (DS-iSYS 25-hydroxvitamin D, Immunodiagnostic Systems, UK). Participants with serum 25-OHD levels <50nmol/L were classified as “deficient” ([54-56], reviewed in [2, 57]). Please note that there is no consensus on the optimal vitamin D level and numerous definitions of vitamin D “deficiency”, “insufficiency” and “inadequacy” exist [2].

Vitamin D supplementation was determined by searching the participants individual medication lists for the ATC-code for vitamin D and its analogs (“A11CC”). Additionally, participants were specifically asked for vitamin D supplementation at a different point during the examination. Only participants who gave consistent answers are kept in the supplemented (“treated”) group and cases with inconsistent answers were ignored during the group matching process.

### Quasi-interventional study design

A quasi-interventional study design was employed to analyze the effect of vitamin D supplementation on DNAmAA. Inclusion criteria for the “treated” group were: (i) vitamin D deficient at baseline, (ii) no vitamin D supplementation at baseline, (iii) start of vitamin D supplementation during follow-up period, (iv) sufficient vitamin D level at follow-up. To identify valid groups for comparison [58], optimal pair matching was performed based on chronological age, sex and a modified version of Charlson’s morbidity index [59, 60]. It was executed with R 3.6.2 [61] and the “MatchIt” package [62] which relies on functions from the “optmatch” package [63]. The “optimal pair matching” method selects matched samples with the smallest mean of the absolute pair distance across all pairs. It is very similar to the often used “nearest neighbor” method but reduces the distance within each pair better [64]. Absolute standardized mean differences before and after the matching process are displayed in Supplementary Figure 3 and 5. Both comparison groups (“untreated” and “healthy”) were determined according to the same procedure.

### Covariates

We included sex and chronological age (years since birth) as covariates in the regression analyses and the group matching process. To be able to adjust for seasonal changes in vitamin D level, we defined the period between October and April as “winter”. This seasonal stratification is in consistency with the analyses at baseline examination [36].

### Statistical Analyses

All statistical analyses and figures were produced with R 3.6.2 [61] and the packages “ggplot” [65] and “ggalluvial” [66]. Linear regression analyses were performed with the “lm” function. We defined participants whose DNAm age parameters differed more than 3 SD from the mean as outliers. The regarding cut-off values were calculated for each individual epigenetic clock parameter considered in this study. A p-value below 0.05 was considered statistically significant.

## Results

### Study population and descriptive statistics

The analyzed longitudinal cohort consists of 1,036 participants of BASE-II that provided information on their vitamin D status at baseline examination (mean age: 68.3 years, SD = 3.5 years) and at reexamination on average 7.4 years later (as part of the GendAge study, mean age: 75.6 years, SD = 3.8 years). Cohort characteristics are displayed in Table 1 and sex-stratified information is available in Supplementary Table 1 and 2.

**Table 1:**
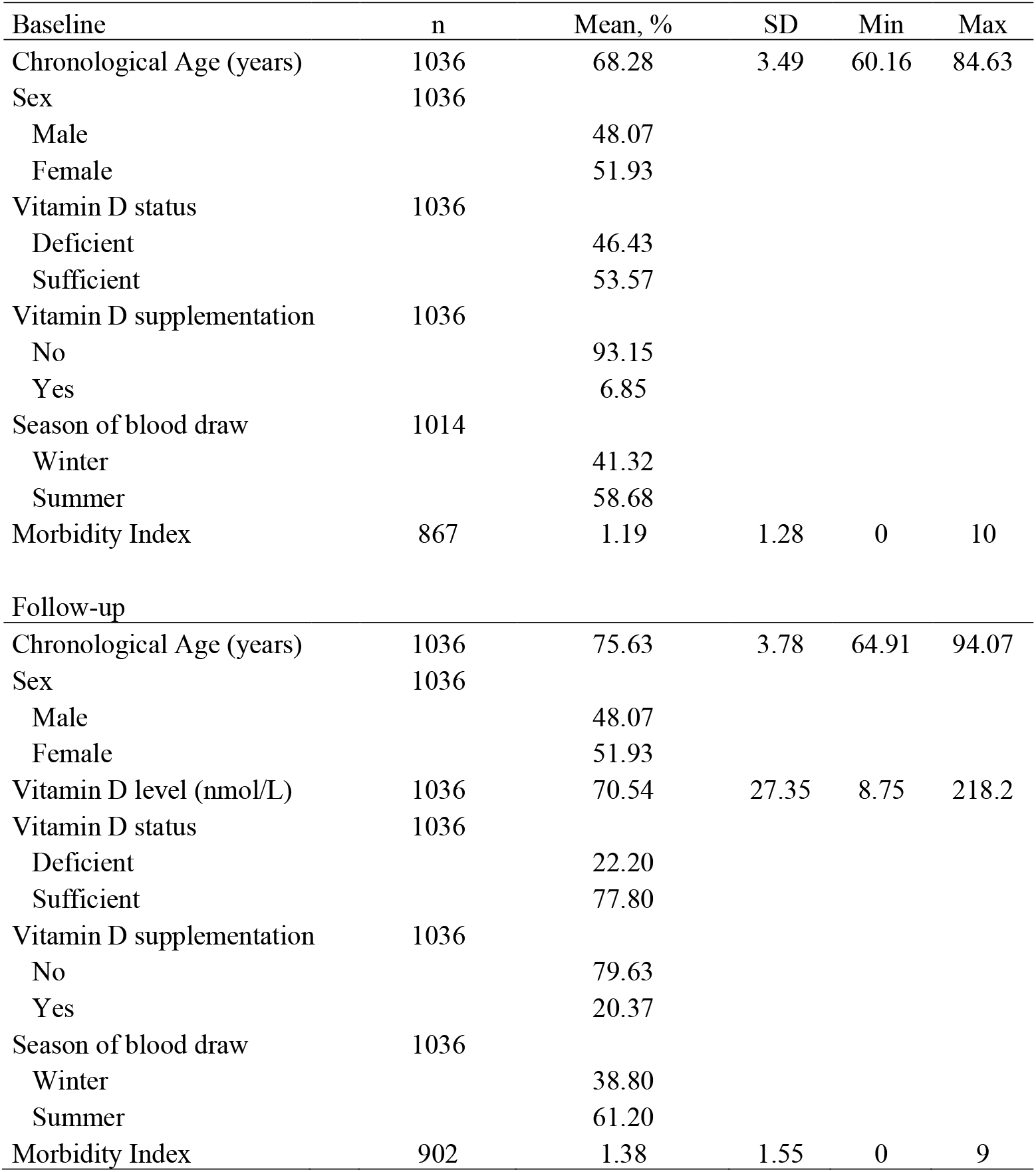
Cohort characteristics of the longitudinally analyzed sample of participants of the BASE-II and GendAge study.

| Baseline | n | Mean, % | SD | Min | Max |
| --- | --- | --- | --- | --- | --- |
| Chronological Age (years) | 1036 | 68.28 | 3.49 | 60.16 | 84.63 |
| Sex | 1036 |  |  |  |  |
| Male |  | 48.07 |  |  |  |
| Female |  | 51.93 |  |  |  |
| Vitamin D status | 1036 |  |  |  |  |
| Deficient |  | 46.43 |  |  |  |
| Sufficient |  | 53.57 |  |  |  |
| Vitamin D supplementation | 1036 |  |  |  |  |
| No |  | 93.15 |  |  |  |
| Yes |  | 6.85 |  |  |  |
| Season of blood draw | 1014 |  |  |  |  |
| Winter |  | 41.32 |  |  |  |
| Summer |  | 58.68 |  |  |  |
| Morbidity Index | 867 | 1.19 | 1.28 | 0 | 10 |
| Follow-up |  |  |  |  |  |
| Chronological Age (years) | 1036 | 75.63 | 3.78 | 64.91 | 94.07 |
| Sex | 1036 |  |  |  |  |
| Male |  | 48.07 |  |  |  |
| Female |  | 51.93 |  |  |  |
| Vitamin D level (nmol/L) | 1036 | 70.54 | 27.35 | 8.75 | 218.2 |
| Vitamin D status | 1036 |  |  |  |  |
| Deficient |  | 22.20 |  |  |  |
| Sufficient |  | 77.80 |  |  |  |
| Vitamin D supplementation | 1036 |  |  |  |  |
| No |  | 79.63 |  |  |  |
| Yes |  | 20.37 |  |  |  |
| Season of blood draw | 1036 |  |  |  |  |
| Winter |  | 38.80 |  |  |  |
| Summer |  | 61.20 |  |  |  |
| Morbidity Index | 902 | 1.38 | 1.55 | 0 | 9 |

DNAm age estimation differed between 0.01 years (GrimAge) and 14.4 years (Levine’s clock) from actual chronological age (Table 2). Correlation between DNAm age and chronological age was between Pearson’s r = 0.3 and Pearson’s r = 0.6. The highest correlation between clocks was found for Horvath’s and Hannum’s clock (Pearson’s r = 0.59 [67]). The test-retest correlation of the longitudinally available 7-CpG clock was in this cohort previously shown to be high (Pearson’s r=0.81) [67]. Participants aged epigenetically slower compared to their chronological age (mean slope of change = 0.75, SD = 0.64, [67]). A detailed cross-sectional and longitudinal description of the included epigenetic clocks can be found in ref. [67].

**Table 2:** Descriptive Statistics of the Five Available Epigenetic Clocks at follow-up. DNAm age acceleration (DNAmAA) was calculated in years as residuals of a linear regression of DNAm age on chronological age and leukocyte cell count (neutrophils, monocytes, lymphocytes, eosinophils).

|  | Women and Men |  |  |  |  | Women |  |  |  |  | Men |  |  |  |  | p-value |
| --- | --- | --- | --- | --- | --- | --- | --- | --- | --- | --- | --- | --- | --- | --- | --- | --- |
|  | n | Mean | SD | Min | Max | n | Mean | SD | Min | Max | n | Mean | SD | Min | Max |  |
| Chronological Age | 1083 | 75.62 | 3.77 | 64.91 | 94.07 | 563 | 75.74 | 3.52 | 66.41 | 94.07 | 520 | 75.49 | 4.02 | 64.91 | 90.03 | 0.259 |
| 7-CpG DNAm age | 1071 | 71.14 | 6.82 | 49.45 | 95.49 | 559 | 70.00 | 6.62 | 52.39 | 95.49 | 512 | 72.39 | 6.82 | 49.45 | 95.29 | <0.001 |
| Horvath's DNAm age | 1070 | 72.43 | 4.52 | 59.86 | 92.11 | 560 | 71.89 | 4.28 | 59.86 | 88.17 | 510 | 73.02 | 4.70 | 61.43 | 92.11 | <0.001 |
| Hannum's DNAm age | 1071 | 63.20 | 4.72 | 50.72 | 82.78 | 561 | 62.45 | 4.56 | 50.72 | 82.79 | 510 | 64.03 | 4.76 | 51.44 | 79.83 | <0.001 |
| PhenoAge DNAm age | 1071 | 61.20 | 6.26 | 42.18 | 84.43 | 560 | 60.50 | 6.05 | 43.75 | 84.43 | 511 | 61.95 | 6.41 | 42.18 | 84.04 | <0.001 |
| GrimAge DNAm age | 1072 | 75.63 | 4.52 | 63.14 | 93.13 | 561 | 74.17 | 3.99 | 63.14 | 89.93 | 511 | 77.23 | 4.53 | 63.99 | 93.13 | <0.001 |
| 7-CpG DNAmAA | 1044 | -0.15 | 5.95 | -17.91 | 17.43 | 545 | -1.16 | 5.97 | -17.54 | 16.66 | 499 | 0.97 | 5.73 | -17.91 | 17.43 | <0.001 |
| Horvath DNAmAA | 1043 | -0.03 | 3.81 | -10.40 | 11.53 | 547 | -0.38 | 3.81 | -10.40 | 11.36 | 496 | 0.36 | 3.78 | -8.94 | 11.53 | 0.002 |
| Hannum DNAmAA | 1041 | -0.12 | 3.60 | -10.80 | 10.85 | 545 | -0.79 | 3.50 | -10.80 | 10.85 | 496 | 0.61 | 3.58 | -9.32 | 10.80 | <0.001 |
| PhenoAge DNAmAA | 1044 | -0.09 | 5.16 | -14.74 | 15.82 | 545 | -0.60 | 5.09 | -14.74 | 15.82 | 499 | 0.47 | 5.19 | -13.51 | 15.38 | 0.001 |
| GrimAge DNAmAA | 1039 | -0.05 | 3.19 | -10.03 | 9.91 | 546 | -1.27 | 2.85 | -10.03 | 9.40 | 493 | 1.30 | 3.01 | -8.17 | 9.91 | <0.001 |

Note: DNAm age = DNA methylation age, DNAmAA = DNAm age acceleration.

Of all longitudinally available participants, 46.4% had deficient vitamin D levels and 6.9% reported intake of vitamin D supplements at baseline. At follow-up we found 22.2% of all participants to be vitamin D deficient and the proportion of participants that used supplements increased by 13.5 percentage points to 20.4% (Table 1, Figure 1). This might be explained by a high fraction of participants that was examined during summer. Only for 55 participants the transition from deficient (at T0) to sufficient (at T1) vitamin D levels cannot be explained by supplementation or season of blood draw (Supplementary Figure 1).

**Figure 1:**
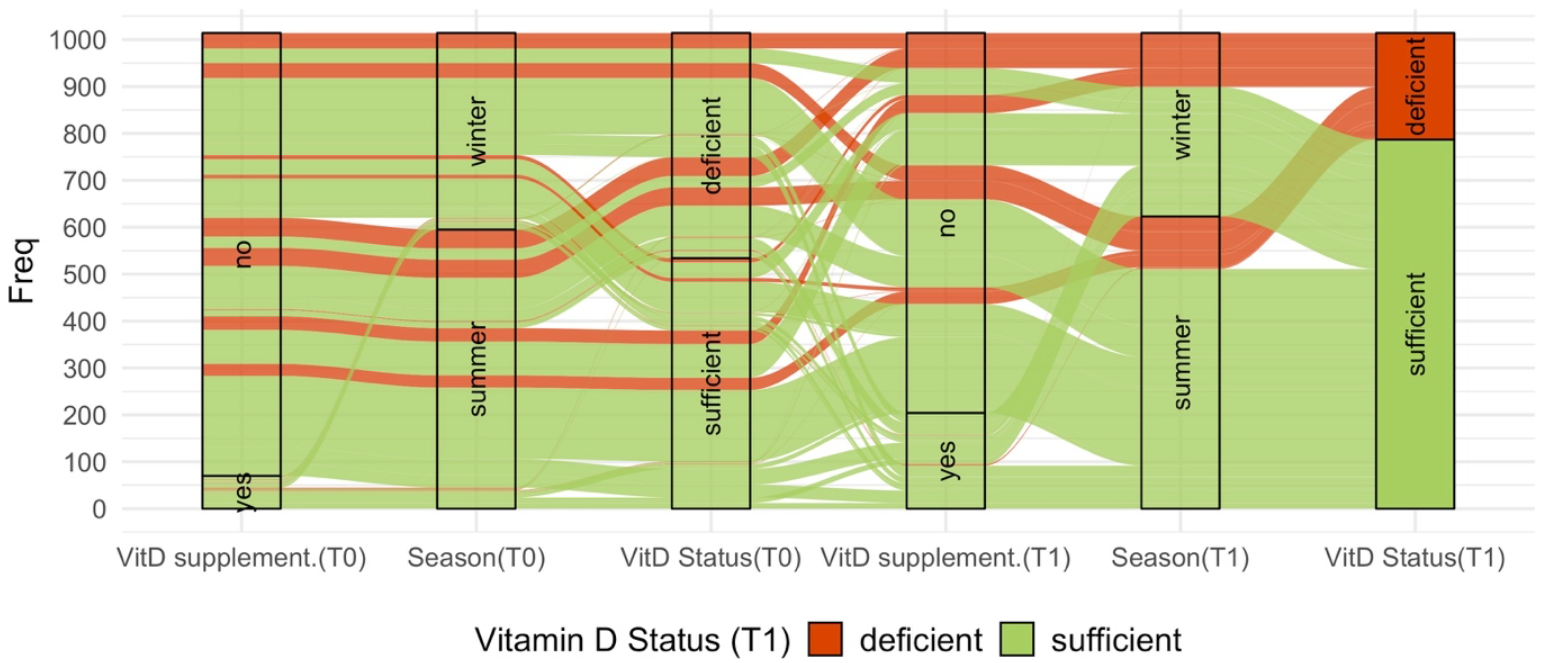
Alluvial plot of vitamin D status and information on supplementation and season of blood draw. The individual trajectories of the 1,036 participants with longitudinal vitamin D data are displayed. Compared to baseline examination, 140 more participants reported intake of vitamin D supplements at follow-up. Note: VitD supplement. = vitamin D supplementation, season = season of blood draw, T0 = baseline examination, T1 = follow-up examination

Of the 71 participants that reported vitamin D supplementation at baseline, 32 stopped within the follow up period. Therefore, 82% of the participants that reported vitamin D supplementation at follow-up started it after baseline examination. The deviation of detected vitamin D deficiency over calendar months during the follow-up examination period showed the expected higher frequency in winter (Supplementary Figure 2).

### Quasi-interventional study: Vitamin D supplementation is associated with lower DNAmAA

We previously reported on a 0.9 year higher 7-CpG DNAmAA in vitamin D deficient BASE-II participants compared to participants with a sufficient vitamin D level at baseline examination [36]. To assess whether this difference persisted after successful treatment of vitamin D deficiency, we employed a quasi-interventional analysis scheme in which the intervention (vitamin D supplementation) was non-randomly assigned by means of self-selection [37]. We compared vitamin D deficient participants that started vitamin D supplementation during follow-up time (and successfully treated vitamin D deficiency as indicated by sufficient serum vitamin level at follow-up) with those who did not and remained with deficient vitamin D level at follow-up. Sixty-three participants met the criteria to be included in the “treated” group (Figure 2 A and B). To minimize selection bias, optimal pair matching was used to select a control group of vitamin D deficient participants without vitamin D supplementation based on chronological age, sex, and morbidity index (Figure 2 A, Supplementary Figure 3).

**Figure 2:**
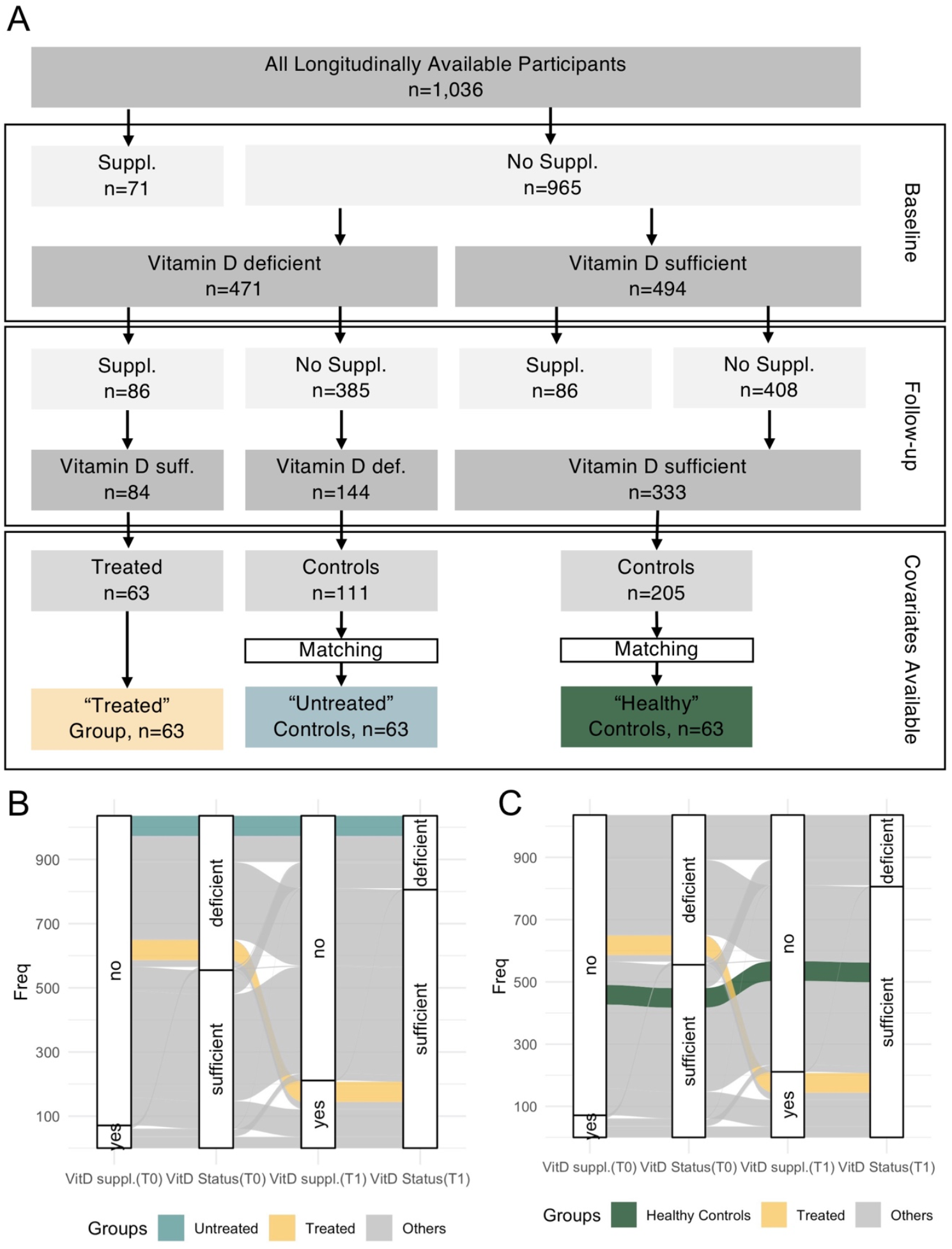
Overview on the longitudinal cohort (A) and of inclusion criteria for the “treated” group (B) and the “untreated” and “healthy” control group (B). VitD suppl. = Vitamin D Supplementation; Supp. = Supplementation., suff. = sufficient, def. = deficient.

We found participants in the treated group to have 2.61 years lower 7-CpG DNAmAA (*t-test*, p=0.011) and 1.27 years lower Horvath DNAmAA (*t-test*, p=0.042) than the participants of the control group of untreated vitamin D deficient participants (Figure 3 B, Supplementary Figure 4). The association between vitamin D supplementation and 7-CpG DNAmAA remained significant after adjustment for covariates (season of blood draw, chronological age, sex) in a multiple linear regression analysis (β = −2.55, SE = 0.99, p = 0.011, n = 126, Supplementary Table 3). Sex-stratified subgroup analyses showed significant associations between 7-CpG DNAmAA and vitamin D supplementation in women (n=86) and between Horvath’s DNAmAA and vitamin D supplementation in men (n=40, Supplementary Table 3).

### Quasi-interventional study: participants with treated vitamin D deficiency do not differ in DNAmAA from vitamin D sufficient participants

We compared participants with successfully treated vitamin D deficiency with participants that reached vitamin D sufficiency without any supplementation at both baseline and follow-up (Figure 2 A and C). These participants were used as “healthy” control group to assess whether the vitamin D deficiency associated accelerated DNAmAA at baseline would still be present after treatment. The same matching process (Supplementary Figure 5) that we described above was employed to select the “healthy” control group (Figure 2 A). No significant difference in DNAmAA was found between participants with treated vitamin D deficiency and the healthy control group (Table 3).

**Table 3:**
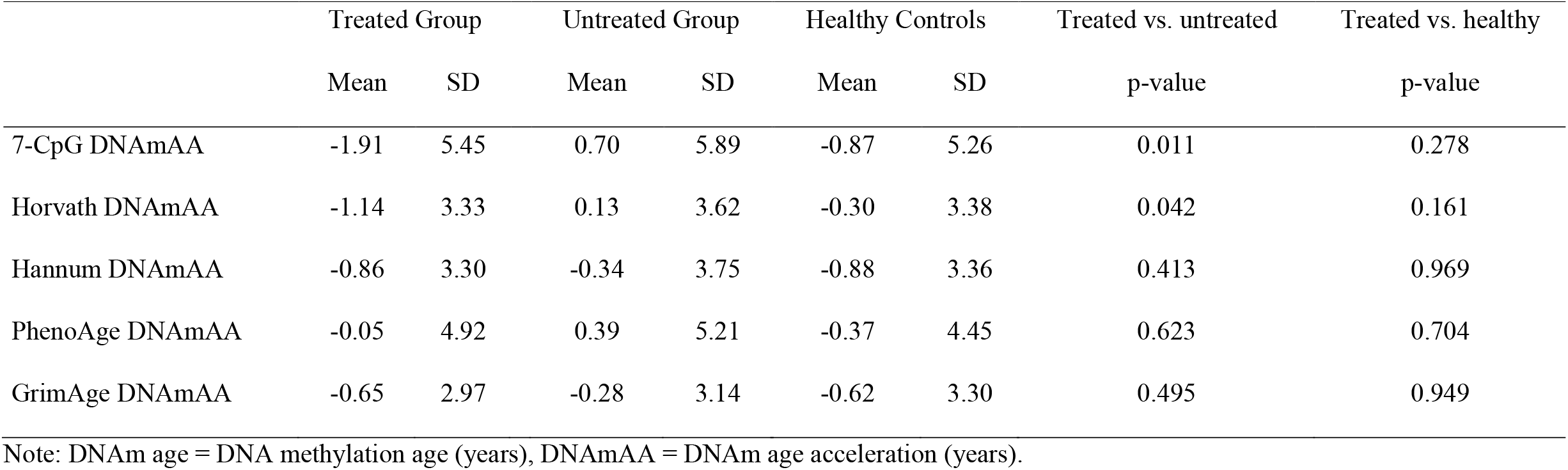
Comparison between participants with successfully treated vitamin D deficiency (“treated”), untreated vitamin D deficient participants and healthy controls.

|  | Treated Group |  | Untreated Group |  | Healthy Controls |  | Treated vs. untreated | Treated vs. healthy |
| --- | --- | --- | --- | --- | --- | --- | --- | --- |
|  | Mean | SD | Mean | SD | Mean | SD | p-value | p-value |
| 7-CpG DNAmAA | -1.91 | 5.45 | 0.70 | 5.89 | -0.87 | 5.26 | 0.011 | 0.278 |
| Horvath DNAmAA | -1.14 | 3.33 | 0.13 | 3.62 | -0.30 | 3.38 | 0.042 | 0.161 |
| Hannum DNAmAA | -0.86 | 3.30 | -0.34 | 3.75 | -0.88 | 3.36 | 0.413 | 0.969 |
| PhenoAge DNAmAA | -0.05 | 4.92 | 0.39 | 5.21 | -0.37 | 4.45 | 0.623 | 0.704 |
| GrimAge DNAmAA | -0.65 | 2.97 | -0.28 | 3.14 | -0.62 | 3.30 | 0.495 | 0.949 |

Note: DNAm age = DNA methylation age (years), DNAmAA = DNAm age acceleration (years).

### Cross-sectional association between vitamin D level and DNAmAA at follow-up

All participants who provided data on their vitamin D level at follow-up were included in the cross-sectional analyses. Too high serum vitamin D levels are harmful [68] and u-shaped association between vitamin D level and health parameters [69, 70] were reported before. Therefore, we excluded 34 participants with potentially harmful vitamin D levels of 125 nmol/L or above [68] from the cross-sectional regression analyses. Individual linear regression analyses of the DNAmAA of five different epigenetic clocks on serum vitamin D level and covariates (season of blood draw, chronological age, sex) were conducted.

We found a significant association between higher levels of vitamin D and 7-CpG DNAmAA (β = −0.02, SE = 0.01, p<0.05, n=1,008) and GrimAge DNAmAA (β = −0.01, SE = 0.004, p=0.03, Table 4). This associations seems to be mainly driven by women, as the all-women subgroup revealed these associations in sex-stratified analyses as well (Table 4).

**Table 4:** Multiple Linear Regression Analysis of DNAmAA of five different epigenetic clocks on Vitamin D level (nmol/L). The model is adjusted for season of blood draw (winter/summer), chronological age (years), and sex (if applicable). Only participants with serum vitamin D levels below 125 nmol/L are included in these analyses.

| Men and Women | Estimate | SE | p-value |  | n |
| --- | --- | --- | --- | --- | --- |
| 7-CpG DNAmAA | -0.016 | 0.008 | 0.049 | * | 1008 |
| Horvath DNAmAA | 0.004 | 0.005 | 0.413 |  | 1007 |
| Hannum DNAmAA | -0.003 | 0.005 | 0.498 |  | 1005 |
| PhenoAge DNAmAA | -0.003 | 0.007 | 0.624 |  | 1008 |
| GrimAge DNAmAA | -0.008 | 0.004 | 0.034 | * | 1005 |
| Men |  |  |  |  |  |
| 7-CpG DNAmAA | -0.004 | 0.012 | 0.705 |  | 485 |
| Horvath DNAmAA | 0.004 | 0.008 | 0.620 |  | 482 |
| Hannum DNAmAA | -0.002 | 0.007 | 0.742 |  | 482 |
| PhenoAge DNAmAA | 0.006 | 0.011 | 0.559 |  | 485 |
| GrimAge DNAmAA | -0.003 | 0.006 | 0.584 |  | 480 |
| Women |  |  |  |  |  |
| 7-CpG DNAmAA | -0.026 | 0.011 | 0.021 | * | 523 |
| Horvath DNAmAA | 0.004 | 0.007 | 0.527 |  | 525 |
| Hannum DNAmAA | -0.004 | 0.007 | 0.548 |  | 523 |
| PhenoAge DNAmAA | -0.012 | 0.009 | 0.222 |  | 523 |
| GrimAge DNAmAA | -0.013 | 0.005 | 0.017 | * | 525 |

## Discussion

In this study we report on a quasi-interventional analyses scheme that revealed significant associations between lower 7-CpG and Horvath DNAmAA and vitamin D supplementation in vitamin D deficient participants. Participants with successfully treated vitamin D deficiency had on average 2.6 years (7-CpG clock, p=0.011) and 1.3 years (Horvath’s clock, p=0.042) lower DNAmAA compared to the group of participants with untreated vitamin D deficiency. Further, we were able to show, that successfully treated participants do not differ in biological age from participants that have sufficient vitamin D levels.

Although vitamin D supplementation in the context of disease and mortality is subject to an intensive discussion, the body of literature analyzing its relationship to DNAmAA is limited. To our knowledge, only one study is available that employs an interventional design. Chen and colleagues conducted a small randomized controlled trial with 51 overweight and vitamin D deficient African Americans (26.1 ± 9.3 years old) that were assigned to four groups (600 IU/d, 2000 IU/d, 4000 IU/d vitamin D3 and placebo). Participants that took 4000 IU/d for 16-weeks showed 1.85 years decrease in Horvath aging compared to the placebo group (p=0.046). A 1.9-year decrease in Hannum aging was found in the group that took 2000 IU per day (p=0.044) [35]. Two cross-sectional studies analyzing the relationship between DNAmAA and vitamin D are available. Schöttker and colleagues found no cross-sectional association between Horvath’s or Hannum’s clock and vitamin D levels in a large cohort of 9,940 participants between 50 and 74 years [4]. In contrast to these findings, we reported on a cross-sectional association between lower 7-CpG DNAmAA and vitamin D sufficiency before. These cross-sectional findings [36] were repeatable in the same participants at follow-up on average 7.4 years later. The additional analyses with DNAmAA estimated by four epigenetic clocks, that rely on epigenomewide methylation data and are available only for follow-up examination, revealed a cross-sectional association between low GrimAge DNAmAA and high vitamin D level (β = −0.01, SE = 0.004, p=0.03). These associations were found in the all-female but not in the all-male subgroups. Men and women seem to epigenetically age at a different pace which was shown in this study (Table 2 and ref. [67, 71]) as well as others [72-74]. Therefore, sex differences in the association between DNAmAA and vitamin D level seem plausible.

Furthermore, it is known that the epigenetic clocks analyzed here differ in the aspects of aging they reflect [33]. This presumably results from the difference in CpG sites included and whether the epigenetic clocks were trained to predict chronological age (first-generation clocks) or phenotypic measures (“second-generation clocks”). Therefore, differences in association with vitamin D between different clocks are expected and help to further differentiate the areas of aging they represent best.

This study has several limitations we want to point out. First, the participants of this study are above average health and the proportion of vitamin D deficient participants (22.2%) is comparatively low. In a large study of the Robert-Koch Institute (DEGS, n=6,995, age range: 18-79 years), 69.9% of men and 62.6% of women between 65 and 79 years had a vitamin D level below 50 nmol/L [75]. The high proportion of participants with sufficient vitamin D levels at follow-up of our study might in part be explained by the high percentage of vitamin D supplemented participants. The frequency of participants with vitamin D supplementation increased by 13.5 percentage points at baseline to 20.37% at follow-up. This rise might be partly explained by the increasing popularity of vitamin D supplements that are easily accessible and comparatively cheap. At baseline examination each participant was given a medical report that included information on the vitamin D level, which might have added to this effect. Therefore, it would be of great interest to repeat these analyses in sample sets with a higher proportion of vitamin D deficient participants. Second, although the sample size for the cross-sectional analyses is comparatively large (n=1,008), the number of participants that meets the criteria to be included in the “treated” group” is rather small. We therefore cannot rule out that missing statistical significance in our study reflects a lack of power. Third, a limitation of this study is the missing information on the exact doses of vitamin D supplementation and duration of intake.

However, the dose for optimal treatment depends on several factors to be considered such as the initial vitamin D level, time spent in the sun, diet and chronological age and needs to be determined individually and to be adjusted if conditions change. Therefore, we used successful treatment (controlled by serum vitamin D level) rather than a specific dose as definition for the “treated” group. A cautious approach to vitamin D supplementation is important because uncontrolled intake of high doses of vitamin D supplements can lead to severe adverse effects [76]. Vitamin D serum levels above 374 nmol/L are associated with hyperphosphatemia and hypercalcemia [2] which can lead to renal failure and cardiac arrest [57]. The Institute of Medicine reported that a vitamin D serum level above 125 nmol/L over a long time period “should raise concerns among clinicians about potential adverse effects” [68]. Therefore, the supplementation of vitamin D, although at least to some extent undoubtfully useful, should be done attentively and only after medical consultation to avoid excessive (as well as insufficient) treatment. Fourth, no causal conclusion can be drawn from the analyses conducted here because we cannot confidently assume that all possible (known and unknown) covariates were considered. It is likely that some of the participants who started vitamin D supplementation during the follow-up period also tried to improve their lifestyle in other ways. As these possible lifestyle changes are not known and therefore not controlled for, they could confound the results reported in this study. To minimize potential confounding effects and selection bias we applied optimal pair matching with carefully selected variables to form valid comparison groups [58]. Nevertheless, only randomized controlled trials are able to control for all confounding factors (known or unknown). Fifth, due to the quasi-interventional study design we do not have a placebo control group. Again, this would be needed to fully understand the potential causal effect of vitamin D on DNAmAA but can only be obtained from randomized controlled trials. Sixth, due to the very limited knowledge of the pathways leading to age related epigenetic changes, we do not know whether vitamin D has an actual influence on biological aging or rather just modifies epigenetic signatures and thus interferes with the measurement accuracy of DNAmAA as an aging marker. The current body of literature makes the former most likely [77, 78], but only sufficiently sized randomized controlled trials can clarify this assumption. Seventh, in line with most other studies that evaluate more than one epigenetic clock, we did not adjust our analyses for multiple testing. Therefore, an increase in the rate of false-positive findings cannot be ruled out and future work needs to independently validate our findings before any further reaching conclusions can be drawn.

Strengths of this study include its longitudinal design and the large, well characterized cohort. It allows to test for associations between vitamin D supplementation and DNAmAA in well balanced intervention and control groups. We compare results across five different epigenetic clocks that are known to represent different domains of aging. Therefore, this study helps to distinguish the domains that are represented best by the different versions of DNAmAA and adds valuable information on the consequences of vitamin D deficiency and how it possibly could be reversed.

## Conclusion

We found DNAmAA to be on average 2.6 years (7-CpG clock, p = 0.011) and 1.3 years (Horvath clock, p = 0.042) lower in vitamin D deficient participants who were successfully supplemented compared to vitamin D-deficient participants without supplementation. Additionally, no difference in DNAmAA of all five clocks was found between treated participants and the control group of participants who are vitamin D sufficient without supplementation. Therefore, a sufficient supplementation of vitamin D-deficient patients seems to be beneficial with regards to epigenetic aging, at least when measured with the 7-CpG or Horvath’s clock. Sufficiently sized randomized controlled trials need to further investigate this potentially causal effect.

## Data Availability

Data are available upon reasonable request. Interested investigators are invited to contact the study coordinating PI Ilja Demuth at to obtain additional information about the GendAge study and the data-sharing application form.

## Acknowledgments

-

## Author contributions

Conceived and designed the study: VMV and ID. Contributed study specific data: all authors. Analyzed the data: VMV and YS. Wrote the manuscript: VMV and ID. All authors revised and approved the manuscript.

