## Supplement for "Vitamin D Supplementation is Associated with Slower Epigenetic Aging"

**Supplementary Table 1: Sex-stratified cohort characteristics of the older participants at baseline examination (BASE-II).** The t-test was used to assess the significance of the differences in means for continuous variables. Difference between categorical variables was determined by chi-square and goodness-of-fit test.

| | female | | | | | male | | | | | |
|---|---|---|---|---|---|---|---|---|---|---|---|
| | n | Mean, % | SD | Min | Max | n | Mean, % | SD | Min | Max | p-value |
| Chronological Age | 538 | 68.07 | 3.33 | 61.34 | 84.63 | 498 | 68.50 | 3.65 | 60.16 | 80.65 | 0.046 |
| sex | 538 | | | | | 498 | | | | | |
| male | 0 | 0.00 | | | | 498 | 100.00 | | | | <0.001 |
| female | 538 | 100.00 | | | | 0 | 0.00 | | | | |
| Vitamin D status | 538 | | | | | 498 | | | | | |
| deficient | 253 | 47.03 | | | | 228 | 45.78 | | | | 0.735 |
| sufficient | 285 | 52.97 | | | | 270 | 54.22 | | | | |
| Vitamin D supplementation | 538 | | | | | 498 | | | | | |
| no | 484 | 89.96 | | | | 481 | 96.59 | | | | <0.001 |
| yes | 54 | 10.04 | | | | 17 | 3.41 | | | | |
| Season of blood draw | 516 | | | | | 498 | | | | | |
| winter | 170 | 32.95 | | | | 249 | 50.00 | | | | <0.001 |
| summer | 346 | 67.05 | | | | 249 | 50.00 | | | | |
| Morbidity Index | 433 | 1.11 | 1.21 | 0 | 10 | 434 | 1.28 | 1.34 | 0 | 7 | 0.050 |

**Supplementary Table 2: Sex-stratified cohort characteristics of the older participants at follow-up examination (as part of the GendAge study).** The t-test was used to assess the significance of the differences in means for continuous variables. Difference between categorical variables was determined by chi-square and goodness-of-fit test.

| | female | | | | | male | | | | | |
|---|---|---|---|---|---|---|---|---|---|---|---|
| | n | mean | sd | min | max | n | mean | sd | min | max | p |
| Chronological Age | 538 | 75.75 | 3.55 | 66.41 | 94.07 | 498 | 75.51 | 4.02 | 64.91 | 90.03 | 0.312 |
| sex | 538 | | | | | 498 | | | | | <0.001 |
| male | 0 | 0.00 | | | | 498 | 100.00 | | | | |
| female | 538 | 100.00 | | | | 0 | 0.00 | | | | |
| Vitamin D level (nmol/L) | 538 | 72.82 | 27.78 | 8.75 | 218.20 | 498 | 68.07 | 26.68 | 8.75 | 200.3 | 0.005 |
| Vitamin D status | 538 | | | | | 498 | | | | | 0.555 |
| deficient | 115 | 21.38 | | | | 115 | 23.09 | | | | |
| sufficient | 423 | 78.62 | | | | 383 | 76.91 | | | | |
| Vitamin D supplementation | 538 | | | | | 498 | | | | | <0.001 |
| no | 387 | 71.93 | | | | 438 | 87.95 | | | | |
| yes | 151 | 28.07 | | | | 60 | 12.05 | | | | |
| Season of blood draw | 538 | | | | | 498 | | | | | 0.824 |
| winter | 211 | 39.22 | | | | 191 | 38.35 | | | | |
| summer | 327 | 60.78 | | | | 307 | 61.65 | | | | |
| Morbidity Index | 464 | 1.33 | 1.49 | 0 | 9 | 438 | 1.44 | 1.61 | 0 | 9 | 0.282 |

**Supplementary Table 3: Multiple linear regression analyses of DNAm age and DNAmAA on group (treated/control), season of blood draw, chronological age and sex (if applicable).**

| | Estimate | SE | p-value | | n |
|---|---|---|---|---|---|
| Women and Men | | | | | |
| 7-CpG DNAmAA | -2.55 | 0.99 | 0.0112 | * | 126 |
| Horvath DNAmAA | -1.18 | 0.61 | 0.0557 | . | 126 |
| Hannum DNAmAA | -0.46 | 0.62 | 0.4650 | | 126 |
| PhenoAge DNAmAA | -0.32 | 0.88 | 0.7177 | | 126 |
| GrimAge DNAmAA | -0.35 | 0.48 | 0.4659 | | 126 |
| Women | | | | | |
| 7-CpG DNAmAA | -3.12 | 1.28 | 0.0165 | * | 86 |
| Horvath DNAmAA | -0.57 | 0.79 | 0.4667 | | 86 |
| Hannum DNAmAA | -0.03 | 0.80 | 0.9730 | | 86 |
| PhenoAge DNAmAA | -0.34 | 1.12 | 0.7582 | | 86 |
| GrimAge DNAmAA | -0.28 | 0.55 | 0.6117 | | 86 |
| Men | | | | | |
| 7-CpG DNAmAA | -1.49 | 1.48 | 0.3214 | | 40 |
| Horvath DNAmAA | -2.55 | 0.91 | 0.0077 | ** | 40 |
| Hannum DNAmAA | -1.22 | 0.96 | 0.2091 | | 40 |
| PhenoAge DNAmAA | -0.40 | 1.44 | 0.7811 | | 40 |
| GrimAge DNAmAA | -0.35 | 0.97 | 0,7236 | | 40 |

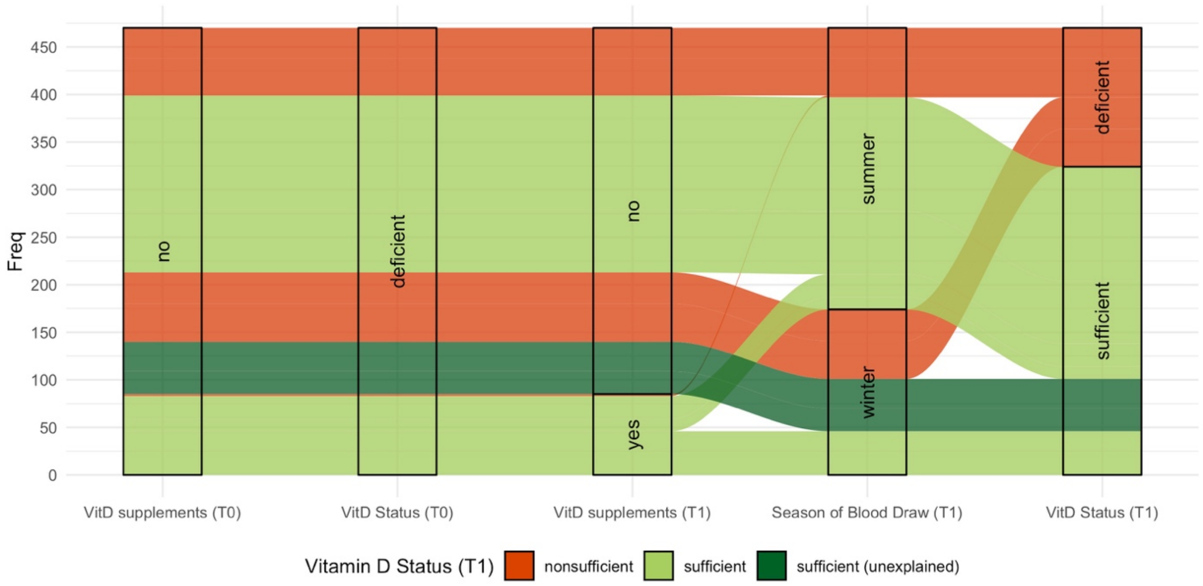


**Supplementary Figure 1: Alluvial plot of participants that showed vitamin deficiency and no vitamin D supplementation at baseline.** Many of the participants that had vitamin D level < 50nmol/L at baseline are vitamin D sufficient at follow-up. This transition can be explained for most participants by either start of vitamin D supplementation or blood draw during summer.

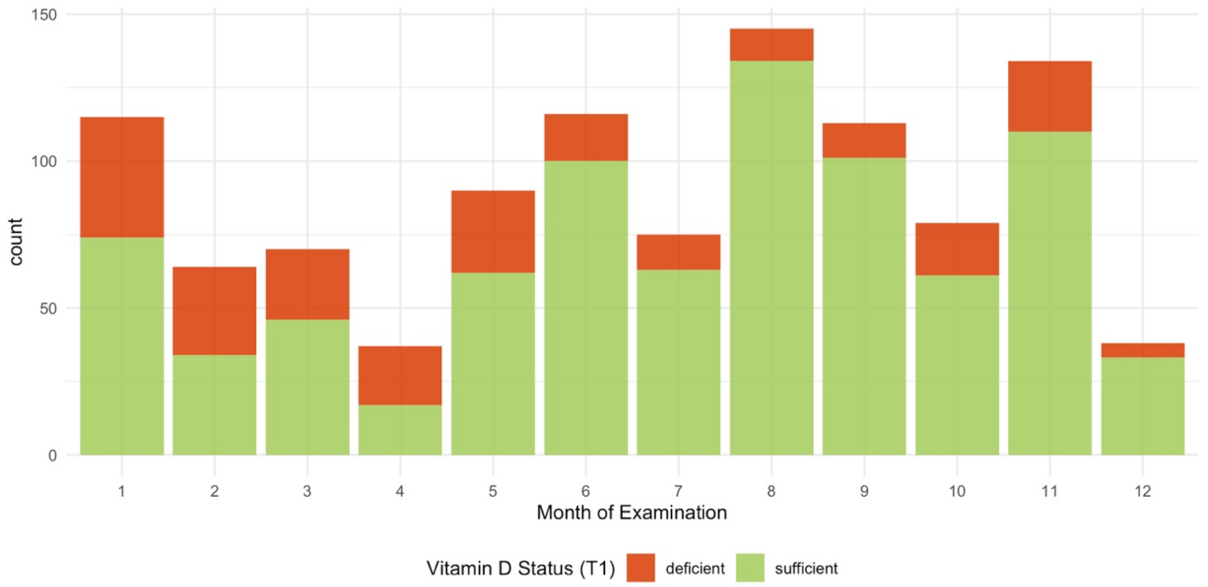


**Supplementary Figure 2: Barplots of the frequency of vitamin D deficient participants stratified by month of blood draw.**

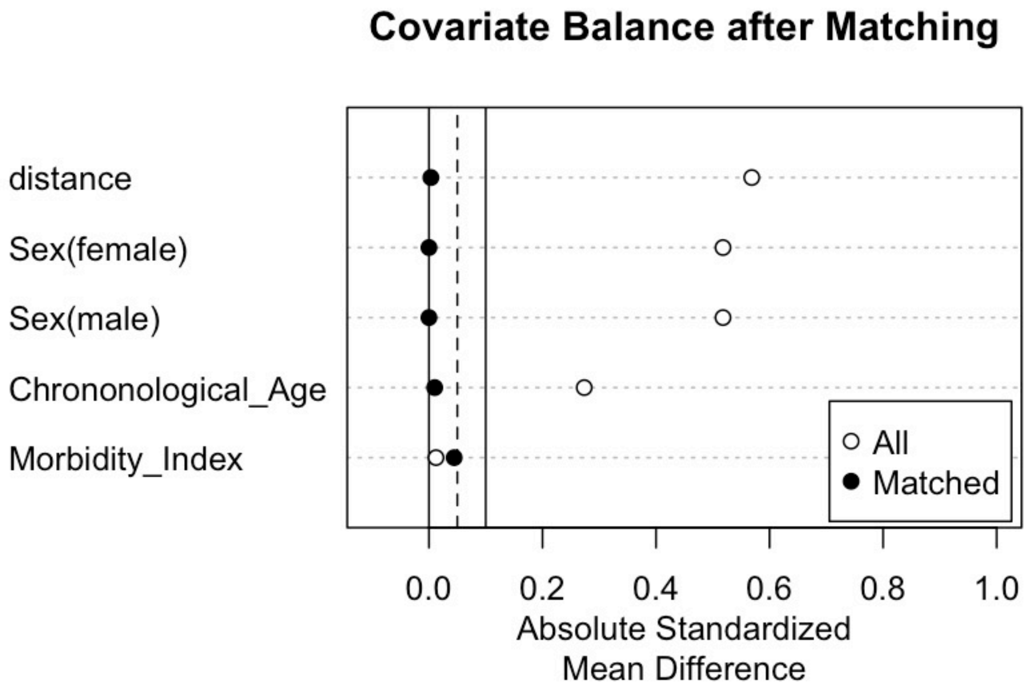


**Supplementary Figure 3: Love plot of absolute standardized mean difference in covariates before and after matching of treated participants with vitamin D deficient participants without supplementation.** Optimal pair matching was performed to identify the ideal control group for the participants that were vitamin D deficient at baseline and started vitamin D supplementation during the follow-up period.

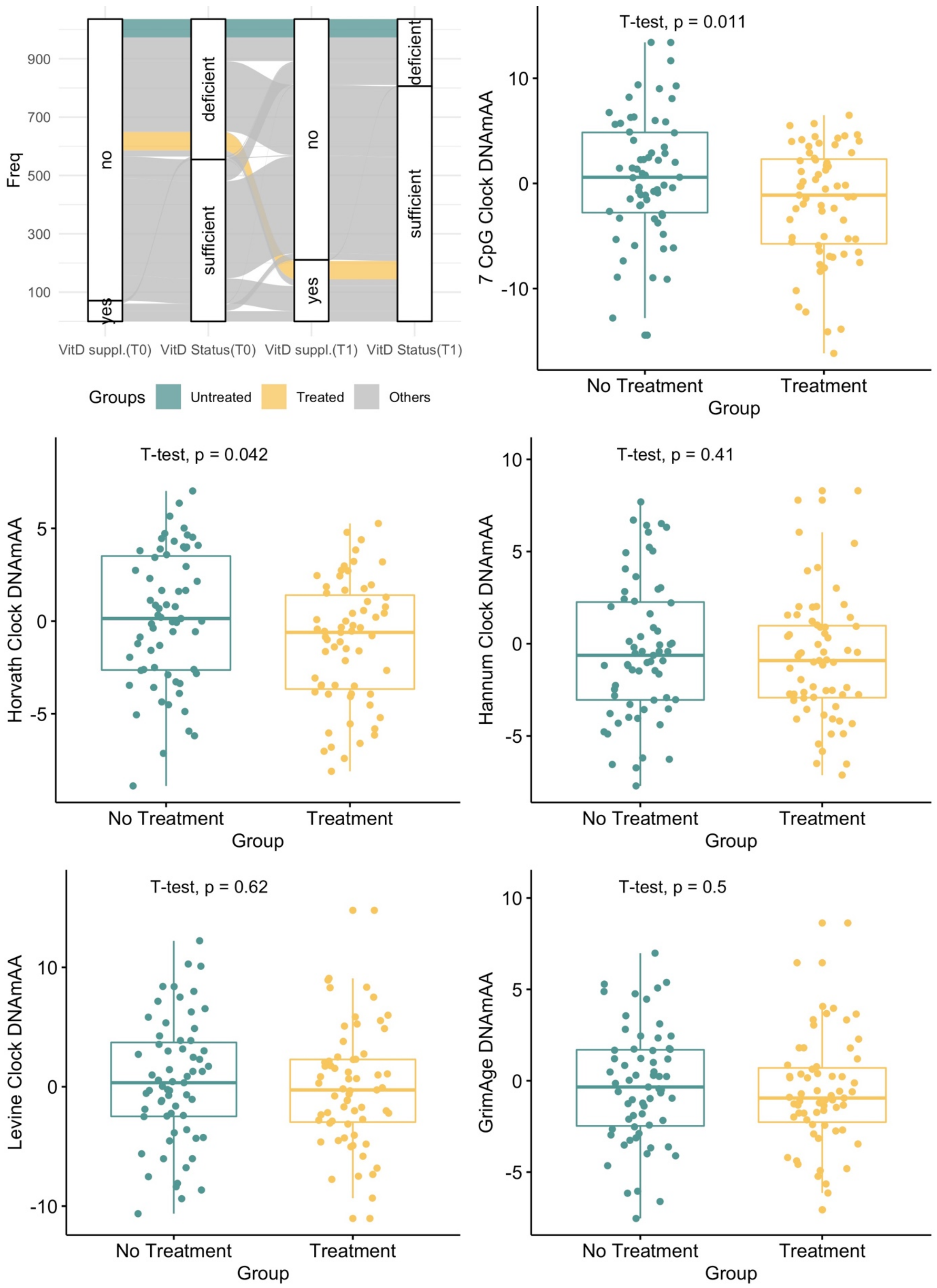


**Supplementary Figure 4: Boxplots of DNAmAA by five different epigenetic clocks in the treated group (n=63) and control group (vitamin D sufficient at baseline and follow-up, n=63).**

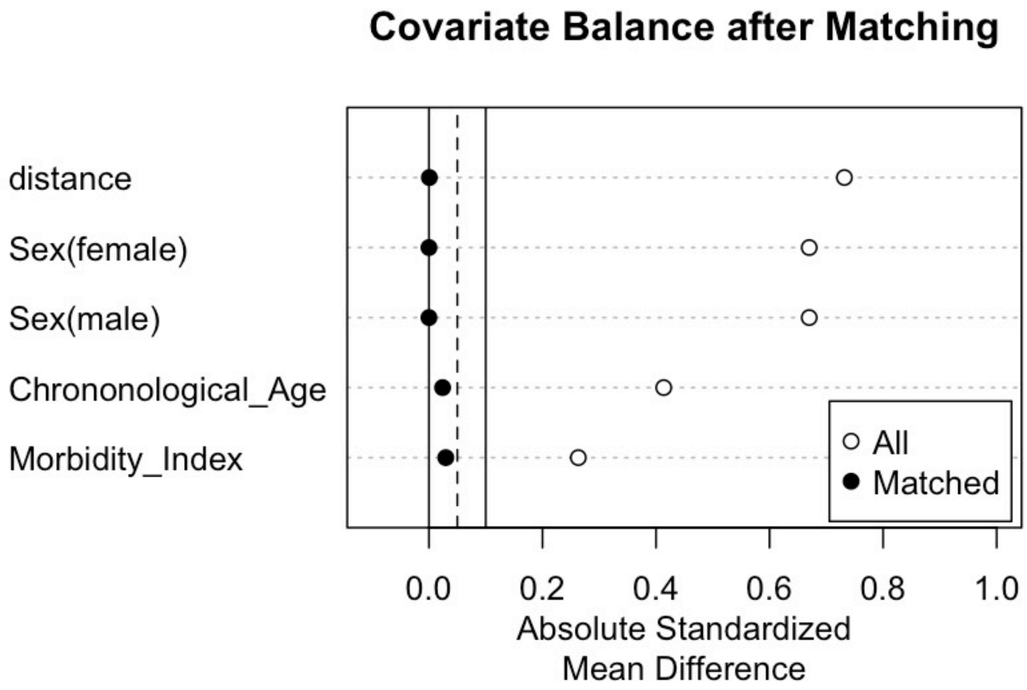


**Supplementary Figure 5: Love plot of absolute standardized mean difference in covariates before and after matching of treated participants with vitamin D sufficient participants without supplementation.** Optimal pair matching was performed to identify the ideal control group of "healthy" participants for the participants that were vitamin D deficient at baseline and started vitamin D supplementation during the follow-up period.
